# Remote Monitoring of AF Recurrence using mHealth Technology (REMOTE-AF)

**DOI:** 10.1101/2023.05.08.23289695

**Authors:** G Adasuriya, A Barsky, I Kralj-Hans, S Mohan, S Gill, Z Chen, J Jarman, D Jones, H Valli, G Gkoutos, V Markides, W Hussain, T Wong, D Kotecha, S Haldar, the REMOTE-AF and CASA-AF trial groups, the cardAIc team and the BigData@Heart Consortium

## Abstract

**Background:** Atrial Fibrillation (AF) detection tools have rapidly developed over the last decade alongside the evolution of mobile health (mHealth) monitoring. mHealth wearable technologies have been hypothesised to be a potential non-invasive and near continuous modality for long term detection and monitoring of atrial arrhythmias. We conducted a proof-of-concept study to evaluate changes in heart rate obtained from a consumer wearable device and compare against implanted loop recorder (ILR)-detected recurrence of AF and atrial tachycardia (AT) after AF ablation.

**Methods:** REMOTE-AF (Remote Monitoring of AF Recurrence Using mHealth Technology; NCT05037136) was a prospectively designed sub study of the CASA-AF randomised controlled trial (NCT04280042). Participants without a permanent pacemaker had an ILR implanted at their index ablation procedure (catheter vs thoracoscopic) for longstanding persistent AF. Heart rate (HR) and step count were continuously monitored using a wrist-worn wearable device connected to a smartphone. Photoplethysmography (PPG) recorded HR data was pre-processed with noise filtration and episodes at 1 -minute intervals over 30 minutes of HR elevations (Z-score = 2) were compared to corresponding ILR data.

Arrhythmias detected by ILR were validated by an independent cardiac physiologist. The AF Effect on Quality of Life (AFEQT) questionnaire was completed by participants at baseline and at the conclusion of the study.

**Results:** Thirty-five patients were enrolled, with mean age 70.3 +/- 6.8 yrs, 12 (34%) women, and median follow-up 10 months (IQR 8-12 months). ILR analysis revealed 17 out of 35 patients (49%) had recurrence of AF/AT. Compared with ILR recurrence, wearable-derived elevations in HR ≥ 110 beats per minute had a sensitivity of 95.3%, specificity 54.1%, positive predictive value (PPV) 15.8%, negative predictive value (NPV) 99.2% and overall accuracy 57.4%. With PPG recorded HR elevation spikes (non-exercise related), the sensitivity was 87.5%, specificity 62.2%, PPV 39.2%, NPV 92.3% and overall accuracy 64.0% in the entire patient cohort. In the AF/AT recurrence only group, sensitivity was 87.6%, specificity 68.3%, PPV 53.6%, NPV 93.0% and overall accuracy 75.0%.

**Conclusion:** Consumer wearable devices have the potential to contribute to arrhythmia detection after AF ablation, but further work is needed to improve and validate new composite detection algorithms.

*Clinical Perspectives:* What is New?

- We have utilised a new composite of data obtained from wearable devices in a predominantly older cohort of patients > 65 years old to detect AF/AT recurrence in a long-standing persistent AF (LSPAF) patient population who have undergone AF ablation.
- We introduce a novel concept of the ‘spike score’, defined as the rate of change in HR over a consecutive two-minute period to detect AF/AT recurrence.
- This is the first study to have achieved this in a post AF ablation cohort of LSPAF compared to the gold standard ILR. Clinical Implications

- The use of wearable devices to look for recurrence of atrial arrythmias in post ablation cohorts may enable a more rapid time to detection for more timely interventions.
- Data composites recorded from wearables may be used alongside the PPG waveform to further improve accuracy in detecting atrial arrythmias.
- AF/AT recurrence detected via wearable devices and associated smart mobile applications can encourage risk factor modification through lifestyle interventions and improve health literacy.

## Introduction

Atrial Fibrillation (AF) is one of the leading causes of cardiovascular morbidity and mortality, increasing the risk of thromboembolism, heart failure and confers a five times increased risk of stroke.^1^ Over 37 million people globally have a diagnosis of AF with its prevalence expected to double by 2060 leading to a significant economic burden.^2^ Given the anticipated increasing health burden of AF, it is a public heath priority to identify patients with AF and those with atrial arrhythmia recurrence post intervention.^3^ The European Society of Cardiology (ESC) AF 2020 guidelines advocate for an integrated care model to be utilised in the diagnosis and management of AF.^1^ A key component of this model requires the use of digital technology and mobile health (mHealth) solutions, to support both health care professionals (HCPs) and patients in the comprehensive and personalised management of health conditions.^4^

Modern and technologically advanced mHealth devices, specifically wearables, have emerged as a new-age, ubiquitous and increasingly accurate solution to monitor cardiovascular biometrics.^5^ However, there is insufficient evidence to demonstrate their utility in current clinical practice. Many published studies lack quality control and are suboptimal in their design, exposing high risk for both selection and publication bias.^6^ Nonetheless, this shift from physician to patient-driven monitoring using data from personable wearable devices demonstrates potential and provides a unique opportunity to identify clinically significant arrhythmias in post intervention patient cohorts. Many of these wearable devices utilise photoplethysmography (PPG) to monitor heart rate and accelerometers to assist with signal processing to filter noise artefact. PPG leverages an optical technique to passively detect pulsatile volumetric changes in blood volume within the peripheral microcirculation to derive a waveform which correlates with the cardiac cycle.^7, 8^

Hence, artificial intelligence (AI) systems have been programmed into wearable devices to accurately predict and detect atrial arrhythmias from PPG waveforms.^9^

In August 2020, the Heart Rhythm Society (HRS) in conjunction with the European Heart Rhythm Association (EHRA) recognised the potential in the use of mHealth devices in cardiovascular disease evaluation and designated it an important frontier in arrhythmia management.^10^ In addition, the European Society of Cardiology (ESC) has led the way in incorporating the use of single lead ECG diagnosis of AF from mHealth devices into their 2020 guidelines for diagnosis and management of AF.^1^ However, the National Institute of Clinical Excellence (NICE) in the United Kingdom has not yet recommended the use of wearables in their recent AF diagnosis and management guideline (NG196) despite Public Health England (PHE) sponsoring initiatives to promote the use of wearable technologies.^11^

A multitude of research studies have assessed the ability of PPG to screen for asymptomatic or index AF, however there is a paucity of data identifying recurrence of AF post rhythm control intervention. Our study sought to evaluate the correlation between a novel composite of wearable device recorded data with ILR (Reveal LINQ™) detected AF/AT recurrence in a cohort of patients who had undergone an AF ablation procedure for LSPAF.

## Methods

### Study Design

The REMOTE-AF study is a single arm, dual-centre, clinical study (NCT05037136) exploring the validity of PPG recorded heart rate data being used in combination with accelerometer derived step count data to predict the recurrence of atrial arrythmias in a post ablation patient population. We labelled this combination as non-exercise related elevations in HR, defined as a step count < 500 over a continuous 30-minute period. The literature does not define ‘non-exercise’ in relation to step count, however a previous study accepted 100 steps per minute as ‘moderate intensity’ exercise and 2500 per day as a sedentary lifestyle.^12^ The study was approved by the UK NRES ethical review board (20/NI/0089). Participants were recruited from the Long Term Outcomes in CASA-AF study (RfPB NIHR200595, ClinicalTrials.gov: NCT04280042) (Figure 1).^13^ This was an extension of the CASA-AF trial, where 120 adults with symptomatic LSPAF, naïve to previous interventions, EHRA symptom score > 2 and left ventricular ejection fraction ≥40 were recruited between September 2015 and June 2018 and randomised to receive either radiofrequency catheter or thoracoscopic surgical ablation for LSPAF. At the index procedure patients had an ILR (Reveal LINQ™) implanted which provided continuous cardiac monitoring with CARELINK remote monitoring to assess arrhythmia recurrence over an initial 12 month follow up period which was subsequently extended to 36 months.^14^ The ILR had validated detection algorithm for AF/AT.^15^

**Figure 1.**
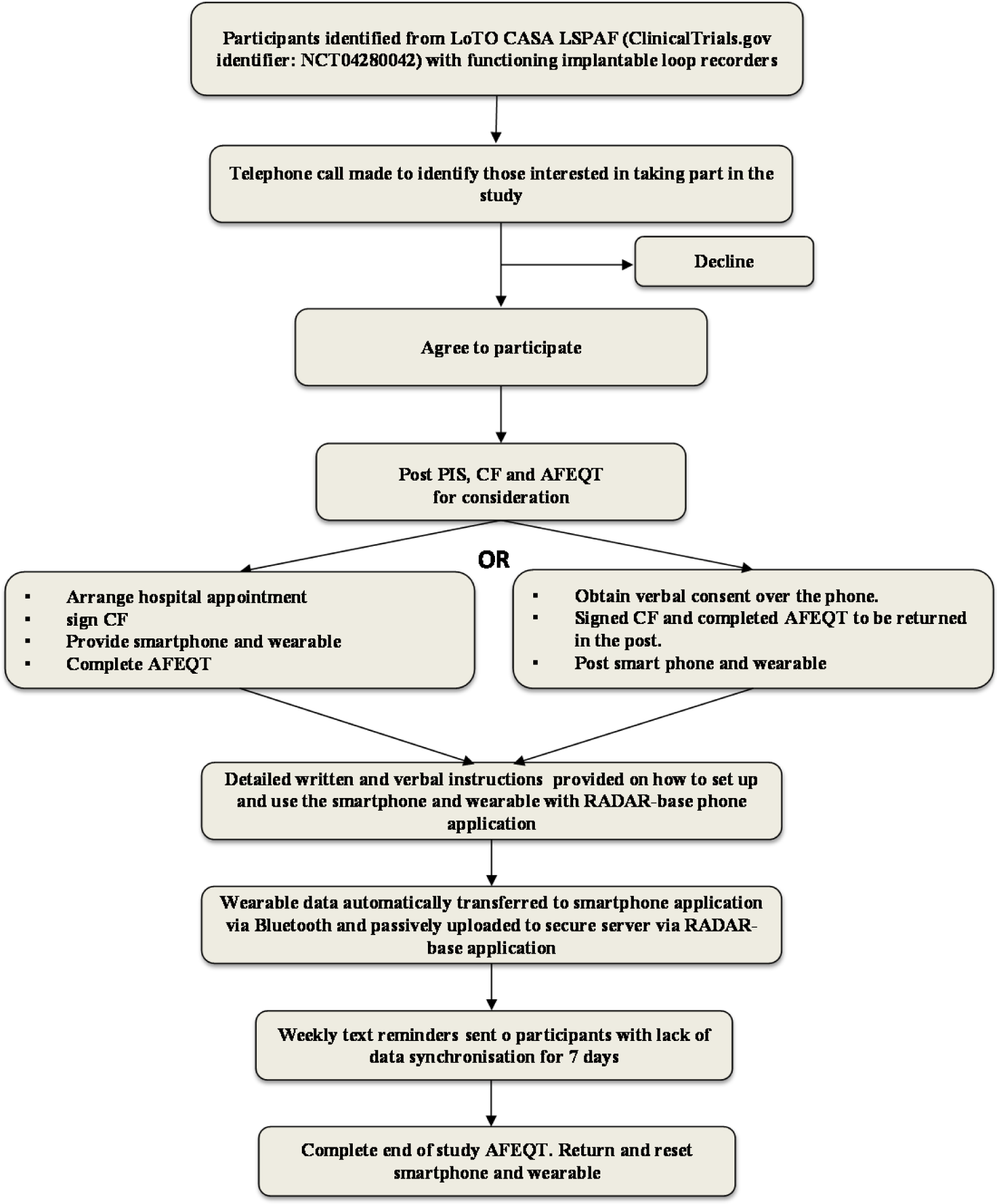
Schematic showing the stepwise process undertaken for patients recruited to REMOTE-AF. LoTO CASA LSAPF = Long-term outcomes in long standing persistent atrial fibrillation. PIS = Patient information sheet. CF = Consent form. AFEQT = AF quality of life survey. RADAR = Remote assessment of diseases and relapses.

### Study Participants

We remotely recruited thirty-five patients, and all provided informed consent. Six eligible participants who met the inclusion criteria (Table 1) declined to take part. Thirty-three patients had ILRs in situ; two patients had dual chamber pacemakers instead of ILRs. Each participant was fitted with a wearable device (Fitbit Charge 2) and provided with a smartphone (Samsung A2) at least 21 months after their index procedure. The remote assessment of disease and relapse (RADAR-base) data collection software and Fitbit mobile application was pre-installed onto the smartphone to allow seamless integration with the Fibit Charge 2 device sensors. The RADAR-base platform streamed data from the wearable, mobile application and smartphone to a central location.^16^ This platform was installed on a virtual machine hosted on Amazon Web Services (AWS) in the AWS Europe (London) region. Researchers were granted access to the Fitbit intraday developer application which allowed automated data collection for registered participants through the RADAR-base platform.

**Table 1:** Inclusion and exclusion criteria for REMOTE-AF. LoTO in CASA LSPAF = Long term outcomes in long standing persistent atrial fibrillation RCT (ClinicalTrials.gov Identifier: NCT04280042). ILR = implantable loop recorder.

| Inclusion Criteria | Exclusion Criteria |
| --- | --- |
| Participants in LoTO in CASA LSPAF with working ILRs or implanted devices capable of continuous rhythm monitoring. | Participants in LoTO in CASA LSPAF without working ILRs or implanted devices capable of continuous rhythm monitoring.<br>Recurrence of Persistent AF.<br>Lack of weekly access to internet or charging facilities. |

The A MEMS 3-axis accelerometer sensor enabled tracking of motion patterns and physical activity whilst the optical wrist-based PPG sensor allowed tracking of heart rate. Participants were encouraged to wear the wearable continuously, removing only during charging and exposure to water, and to connect the Bluetooth enabled smartphone to the internet at least once a day to allow synchronisation of data with the RADAR-base servers. At weekly intervals, participants were reminded via text message to synchronise data if uploads had not occurred and ad-hoc virtual meetings allowed troubleshooting of technical aspects.

Participants were asked to report any adverse effects (skin irritation, health anxiety or stress) related to the use of the wearable. Data was not collected beyond 31^st^ January 2022.

### Data Analysis

Heart rate and step count data recorded by the wearable were transmitted to the smartphone mobile application via Bluetooth. PPG recorded HR data underwent signal processing and was filtered for motion artefact using two accelerometer sensors. The RADAR-base application was programmed to allow access to the Fitbit developer intraday application and subsequently passively collected data and synchronised directly to a secure, encrypted cloud-based web server. At the conclusion of the study, data was downloaded from the cloud server onto a local server for analysis. The data included heart rate and step count time series gathered from wearable devices and pooled at one-minute intervals. Mean heart rate data was calculated for each individual participant at one-minute intervals over the duration of follow up. We developed a method to detect sequences potentially suggestive of atrial arrhythmia recurrence for each patient at thirty-minute overlapping intervals where heart rate was at least two standard deviations (Z score ≥ 2) above their mean for at least twenty minutes and later excluded those that were exercise related. We extracted all of the available tagged thirty-minute sequences for each patient (a total of 3208 sequences across our cohort) and associated within these sequences heart-rate data with step data counterparts at the same time points. Sequences were kept only where heart rate and step-count were both available for the duration of the thirty-minute window. Each of these sequences were then stratified into subcategories where corresponding step count values were denoted at 500 step intervals, with further sub categorisation based on heart rate delta change. We used a simple yet reliable method to identify time points where a participant’s HR spiked. Across each sequence, we searched for the greatest increase in HR across a two-minute interval and recorded the size of that increase (in BPM). We termed this novel composite the “spike score”. To compare across patients, we computed the normalised spike score (or spike z-score), which is this figure divided by the standard deviation of the patient’s heart rate. We also computed the normalised downward spike score for each sequence, which is the largest two-minute decrease in a patient’s heart rate that occurred after the upward spike. Normalised spike scores ≥ 0.75 were taken to be indicators of sudden and significant heart rate elevation. Each data sequence within each subcategory was compared to ILR data to denote if recurrence of an atrial arrythmia had occurred.

For our analysis, a positive finding was recorded when our novel composite method for AF/AT recurrence correlated with ILR detected electrogram (EGM) episodes showing arrhythmia recurrence. If a text only episode of AF/AT was recorded by the ILR, we sought a confirmatory EGM episode within a 24-hour period. ILR detected recurrences of atrial arrhythmia for at least 30 seconds were validated by a blinded senior cardiac physiologist. One patient with no ILR detected AF/AT recurrence provided only three minutes of wearable device data and therefore was excluded from our analysis.

### Study Outcomes

Our primary endpoint is the correlation between a composite of mHealth recorded heart rate (PPG) and physical activity as denoted by step count and AF/AT recurrence as detected by ILR. We analysed this in a stepwise manner:

a. PPG recorded HR elevations (z score ≥ 2) correlating with AF/AT recurrence as detected by ILR.
b. PPG recorded HR elevations (z score ≥ 2) combined with physical activity correlating with AF/AT recurrence as detected by ILR.
c. PPG recorded HR elevations (z score ≥ 2) ≥ 110 beats per minute with normalised spike score ≥ 0.75 combined with physical activity correlating with AF/AT recurrence as detected by ILR.

Our secondary endpoint is the correlation between quality-of-life metrics as measured by the AF quality-of-life questionnaire (AFEQT) score and ILR-detected AF/AT recurrence.

### Statistical Analysis

The sensitivity, specificity, NPV, PPV and accuracy and associated 95% confidence intervals for detection of correlated atrial arrhythmias in individual study participants were calculated using Microsoft Excel (Office 365, Microsoft Corporation, Washington, USA) with a significance level of 0.05. Missing data from the wearable, defined as lack of HR and step count data for the entire 30-minute duration of identified sequences using the method described above, were excluded from the extraction process and not included in final data analysis.

## Results

The mean [SD] age of recruited patients was 70.3 [+/-6.8] years with median 10 months [IQR 8-12] follow up (Table 2). Twenty-five patients completed at least six months follow up with eight of these patients completing the full 15 month follow up. In total, 236,871 hours of data were recorded via the wearable which amounts to a mean [SD] of 282 [+/-3.6] days per participant. On average, participants recorded data during 80.6% of daytime hours (8am and 8pm) and 71.1% of night-time hours (8pm-8am). Implantable loop recorder analysis showed 48.6% (17/35 patients) had recurrence of AF/AT. The average AF Quality of Life Survey (AFEQT)^17^ score for patients in the non-AF/AT recurrence group was 48.9 at the beginning of the study and 32.3 at the end of the follow up period, an improvement of 20% compared to the AF/AT recurrence group who had scores of 45.0 pre study and 40.6 post study.

**Table 2.** Baseline characteristics of REMOTE-AF study participants. AF, atrial fibrillation; AFEQT, AF quality of life survey; BMI, body mass index; CHA_2_DS_2_VASc, congestive heart failure, high blood pressure, Age 75, Diabetes, previous Stroke or clot, Vascular disease, Age 65–74, Sex; HASBLED, hypertension; Abnormal liver/renal function, Stroke history, Bleeding history or predisposition, Labile INR (international normalized ratio), Elderly, Drug/alcohol usage; IQR, interquartile range; SD, standard deviation; TIA, transient ischaemic attack.

|  | All |  |  |
| --- | --- | --- | --- |
| Characteristics | (*n* = 35) | Recurrence (*n* = 17) | No recurrence (*n* = 18) |
| Age (years), mean (SD) | 71 (6.8) | 71 (6.31) | 69 (7.2) |
| ≥ 75, *n* (%) | 10 (28.6) | 5 (29.4) | 5 (27.8) |
| 65-74 | 18 (51.4) | 7 (41.2) | 11 (61.1) |
| 55-64 | 6 (17.1) | 4 (23.5) | 2 (33.3) |
| 40-54 | 1 (2.9) | 1 (5.9) | 0 (0) |
| Male sex, *n* (%) | 23 (65.7) | 9 (52.9) | 14 (77.8) |
| BMI (kg/m<sup>2</sup>), median (IQR) | 30.4 (25.3–33.27) | 26.9 (25–33) | 30.9 (28.5–33.3) |
| Ethnicity, *n* (%) |  |  |  |
| White | 35 (100) | 17 (100) | 18 (100) |
| Ejection fraction (%), mean (SD) | 56.9 (8.9) | 55.2 (8.9) | 58.8 (8.7) |
| Left atrial diameter (mm), mean (SD) | 44.6 (5.9) | 44.6 (6) | 44.7 (5.8) |
| Index procedure, *n* (%) |  |  |  |
| Thoracoscopic surgical ablation | 16 (45.7) | 7 (41.2) | 9 (50) |
| Radiofrequency catheter ablation | 19 (54.3) | 10 (58.8) | 9 (50) |
| Time from index procedure (months), mean (SD) | 34 (7.8) | 33 (5.5) | 37 (6.5) |
| AFEQT score, mean (SD) | 46 (21) | 44 (15) | 48 (25) |
| Medical history, *n* (%) |  |  |  |
| Hypertension | 13 (37.1) | 8 (47.1) | 5 (27.8) |
| Diabetes | 3 (8.6) | 1 (5.9) | 2 (11.1) |
| Coronary artery disease | 2 (5.7) | 1 (5.9) | 1 (5.6) |
| Stroke, TIA, Thromboembolism | 3 (8.6) | 2 (11.8) | 1 (5.6) |
| Heart Failure | 3 (8.6) | 2 (11.8) | 1 (5.6) |
| CHA<sub>2</sub>DS<sub>2</sub>VASc score, *n* (%) |  |  |  |
| 0 | 2 (5.7) | 1 (5.9) | 1 (5.6) |
| 1 | 10 (28.6) | 5 (29.4) | 5 (27.8) |
| 2 | 11 (31.4) | 5 (29.4) | 6 (33.3) |
| ≥3 | 12 (34.3) | 6 (35.3) | 6 (33.3) |
| HASBLED score, *n* (%) |  |  |  |
| 0 | 1 (2.9) | 0 (0) | 1 (5.6) |
| 1 | 9 (25.7) | 6 (35.3) | 3 (16.7) |
| 2 | 22 (62.9) | 10 (58.9) | 12 (66.7) |
| ≥3 | 3 (8.6) | 1 (5.9) | 2 (11.1) |

Analysis of our data in a stepwise manner showed PPG-recorded HR correlating with AF/AT recurrence had a sensitivity of 95.3% (95%CI, 71-99); specificity 54.1% (95%CI, 50-59); PPV 15.8% (95%CI, 4-27); NPV 99.2% (95%CI, 99-100) and accuracy 57.4% (95%CI, 52-63) (Table 3). Combining PPG-recorded heart rate and step count with AF/AT recurrence yielded a sensitivity of 93.2% (95%CI,68–99%); specificity 54.9% (95%CI, 49-60%), PPV 19.1% (95%CI, 6-18%); NPV 98.6% (95%CI, 97-99%) and accuracy 58.7% (95%CI, 52-65%) (Table 3) (Figure 2 and 3). PPG recorded HR sequences identified by a normalised spike score of 0.75 or above over 2 minutes for both onset of heart rate ≥ 110 and cessation below 110 over the same time period produced a sensitivity of 87.5% (95%CI, 63-99%); specificity 62.2% (95%CI, 56-69%); PPV 39.2% (95%CI, 24-54%); NPV 92.3% (95%CI, 89-96%) and accuracy 64.0% (95%CI, 56-71%) (Table 3) (Figure 4). Focused assessment of the latter in the AF/AT recurrence group as defined by ILR led to a sensitivity of 87.6 (95%CI, 65-99%); specificity 68.3% (95%CI, 60-76%); PPV 53.6% (95%CI, 38-69%); NPV 93.0% (95%CI, 88-98%) and accuracy 74.0% (95%CI, 66-82%) (Table 3).

**Figure 2.**
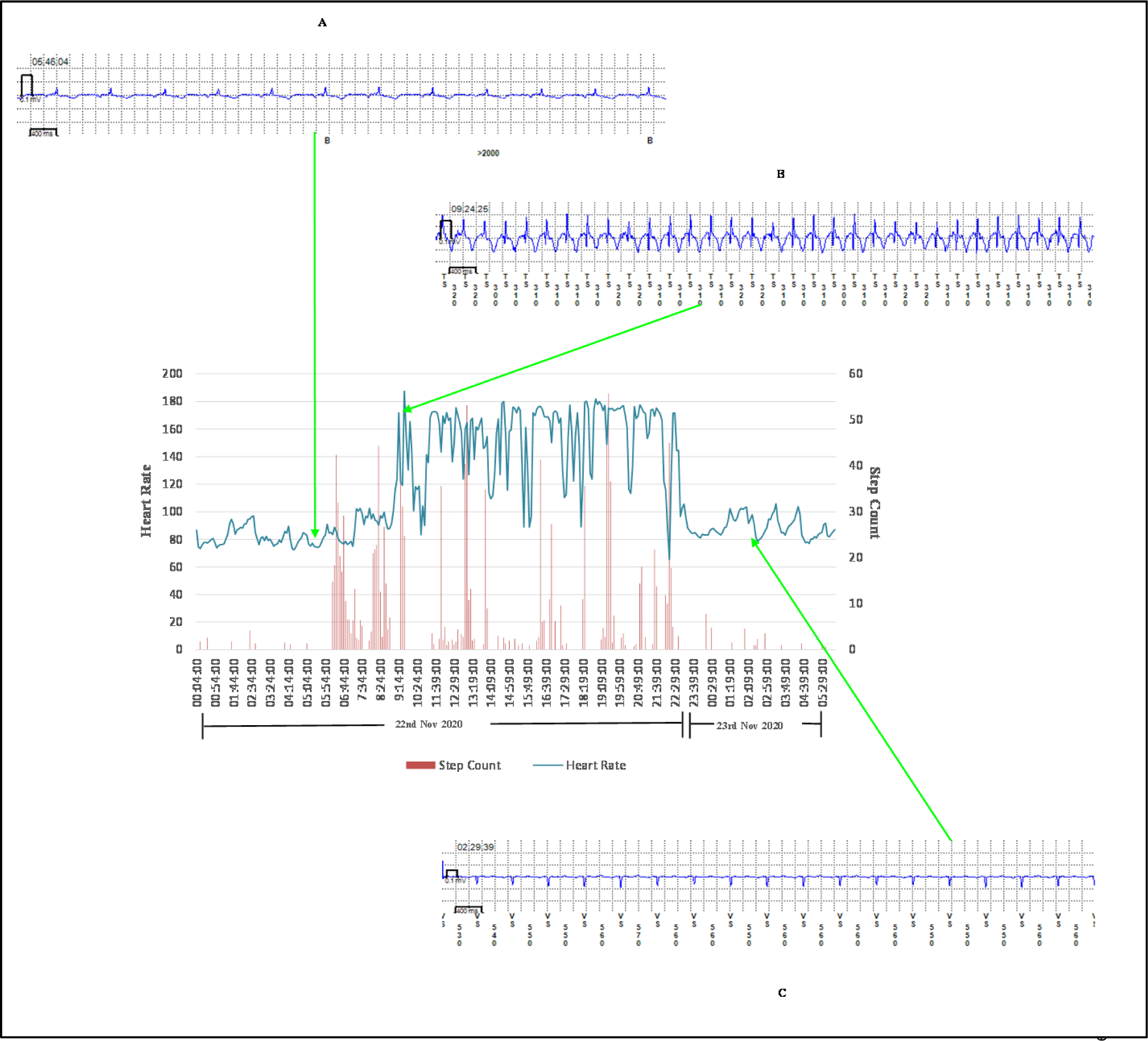
Transition from normal sinus rhythm to atrial tachycardia in a patient with AT recurrence. Heart rate and step count as recorded by a PPG and accelerometer-enabled wrist-worn device demonstrate a period of fixed maximum rate tachycardia and increased beat-to-beat variability, which correlates with ILR-confirmed paroxysmal AT. A. ILR electrogram showing normal sinus rhythm B. ILR electrogram showing atrial tachycardia C. ILR electrogram showing cardioversion back to normal sinus rhythm.

**Figure 3.**
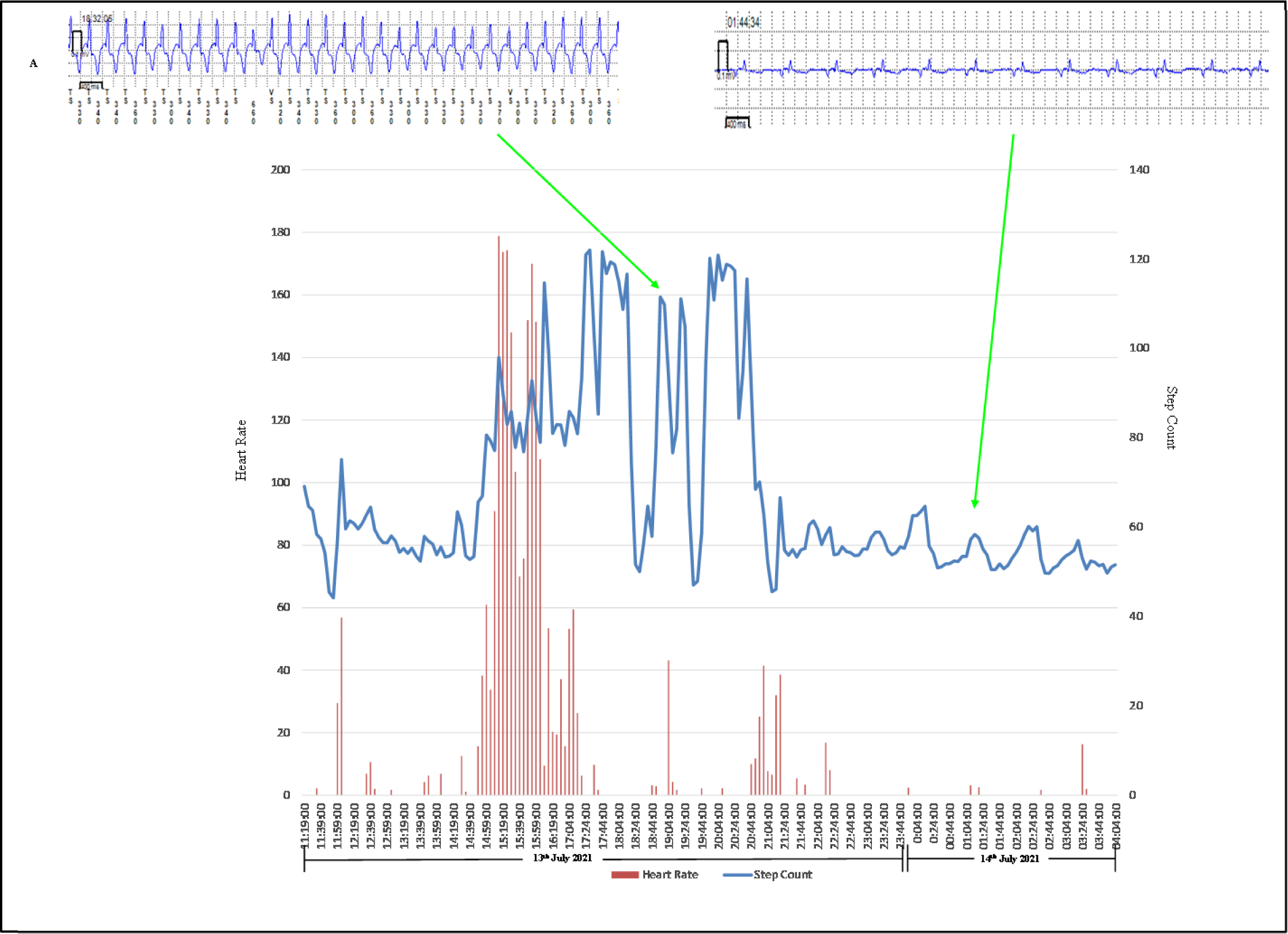
Transition from normal sinus rhythm to atrial fibrillation in a patient with AF recurrence. Heart rate and step count as recorded by a PPG and accelerometer-enabled wrist-worn device demonstrate a period of variable rate tachycardia and increased beat-to-beat variability, which correlates with ILR-confirmed paroxysmal AF. A. ILR electrogram showing atrial fibrillation B. ILR electrogram showing normal sinus rhythm

**Figure 4.**
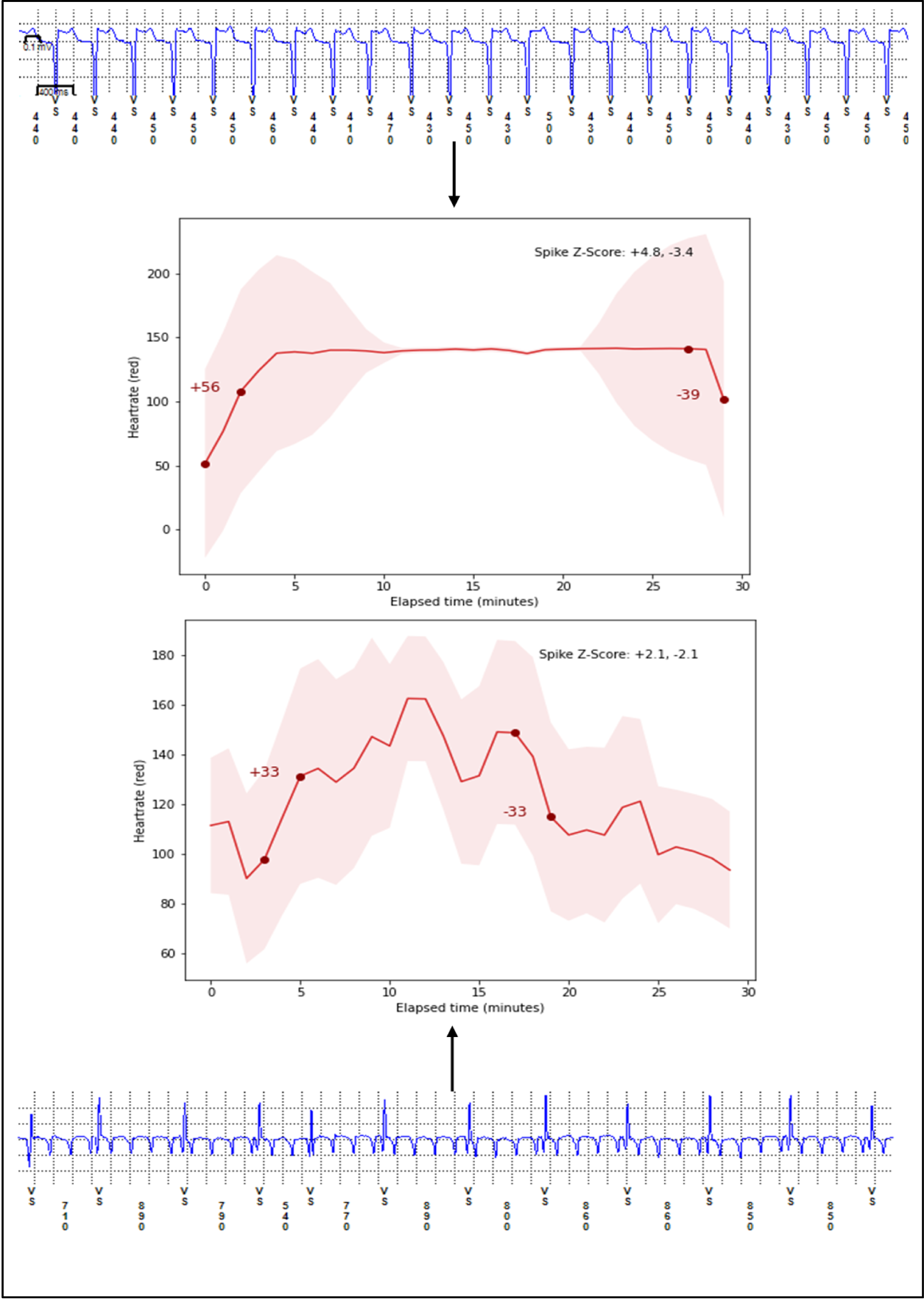
Demonstration of our underlying composite method to detect AF/AT recurrence by utilising matching onset and offset for HR delta change (spike score) and subsequent arrhythmia recurrence as detected by ILR. A normalised spike score ≥ 0.75 for both onset and offset of tachycardia is more likely to represent an episode of atrial arrhythmia recurrence due to a sudden increase in HR rather than exercise induced tachycardia with a gradual increase in HR.

**Table 3:** Results table showing sensitivity, specificity, positive predictive value (PPV), negative predictive value (NPV) and accuracy for each stepwise combination of wearable device data.

|  | Sensitivity<br>(%) | Specificity<br>(%) | PPV<br>(%) | NPV<br>(%) | Accuracy<br>(%) |
| --- | --- | --- | --- | --- | --- |
| Heart Rate | 95.3 | 54.1 | 15.8 | 99.2 | 57.4 |
| Heart Rate and Step Count | 93.2 | 54.9 | 19.1 | 98.6 | 58.7 |
| Heart Rate spike score and Step Count | 87.5 | 62.2 | 39.2 | 92.3 | 64.0 |
| HR spike score and Step Count in recurrence group | 87.6 | 68.3 | 53.6 | 93.0 | 74.0 |

No adverse events such as skin irritation from wearable devices, health anxiety or stress were reported by any of the 35 participants. Three wearable devices required strap replacement and resulted in 9 follow-up days lost.

## Discussion

REMOTE-AF evaluated the correlation between wearable device derived heart rate and AF/AT recurrence. Current physician prescribed non-continuous rhythm monitoring tools (ambulatory Holter monitors) which assess for arrhythmias at set time intervals are known to have low detection yields.^18^ In symptomatic patients with suspected arrhythmias, a 24-hour ambulatory Holter monitor has a diagnostic yield of 7% compared to 47% with a 7-day monitor.^19^ At present, the gold standard for detection of atrial arrhythmias is continuous monitoring via the implantable loop recorder (ILR). mHealth wearable devices can bridge the gap between intermittent time limited Holter monitoring and invasive and costly ILR monitoring.

To analyse our data, we adopted a stepwise approach to sequentially combine parameters from the wearable device. We saw an increase in PPV from 15.8% to 39.2% and improvement in accuracy from 57.4% to 64.0% when using our novel composite HR spike score method compared to using PPG recorded HR alone to detect AF/AT recurrence. This is because, the spike score is more likely to represent true arrhythmia rather than physiological HR increase and hence more accurately differentiate between sinus tachycardia and pathological recurrences of AF/AT. In patients with known AF/AT recurrence as detected by ILR, our novel composite methodology utilising spike score resulted in the highest accuracy in identifying AF/AT recurrence in a post ablation cohort when compared with ILR. Irrespective of sequential improvements in accuracy, using this method results in 5 out of 10 episodes being misclassified as AF/AT. Further work with larger patient populations is needed to improve the diagnostic accuracy of our method. Our results yielded a high NPV and low PPV, likely due to our small sample size with low prevalence of AF/AT recurrence. In clinical practice, this supports the use of wearables in high risk patient populations with known diagnoses of atrial arrhythmias rather than for use in screening in low risk patient populations (global AF prevalence of 0.51%).^2^ Our results also show that our novel composite method in its current form, appropriately classifies non-recurrence of AF/AT but occasionally misclassifies recurrence. Within the current consumer market, the diversity in the accuracy and quality of PPG embedded in wearable devices limits the potential of integrating these data safely into the clinician’s decision-making process.^5, 20^ Extensive research and funding has been dedicated to assessing the accuracy of PPG enabled wearables in detecting atrial arrhythmias, predominantly AF screening but there remains a lack of data in post AF intervention populations. Products developed by multi-billion-dollar global technology giants have shown promise that wearable devices could become a viable alternative in diagnosing and monitoring arrhythmias. The Apple Watch (Apple Inc, Cupertino, CA, USA), Fitbit (Google LLC, Mountain View, CA, USA) and Huawei Smartwatch (Huawei Technologies Co Ltd, Shenzhen, China) have been assessed in non-randomised but large-scale prospective clinical trials, reporting accuracy of 95-97% in detecting AF and modest positive predictive values (PPV).^21–23^

Our statistical analysis revealed a PPV significantly lower (Figure 5) than figures reported by these landmark PPG detected arrhythmia studies, however these large-scale studies were based on AF screening and not recurrence as in our cohort. Based on a recent meta-analysis, these figures are likely to be unrealistically elevated, therefore our results are likely to be more representative of the accuracy of PPG in detecting AF/AT recurrence.^6^ Given there is limited published data, predominantly case reports,^24–27^ documenting wearable use in rhythm monitoring after catheter ablation we are unable to make a direct comparison of PPG enabled wearables in this patient cohort. The reported clinical metric in the landmark studies only analysed PPG HR waveforms intermittently whilst stationary, with the majority at night. We reported results for all data both during active daytime hours and whilst ambulatory, where clinically significant arrhythmias are more likely. Overall, our data provides greater real-world analysis, and highlights the issue of lower diagnostic yields and challenges related to sensitivity and specificity of data from wearable devices during periods of activity. Conducting this study during a global pandemic also produced its own challenges but further advanced the necessity and accessibility for remote monitoring strategies, not just for western populations but worldwide, to ensure health equity.

**Figure 5:**
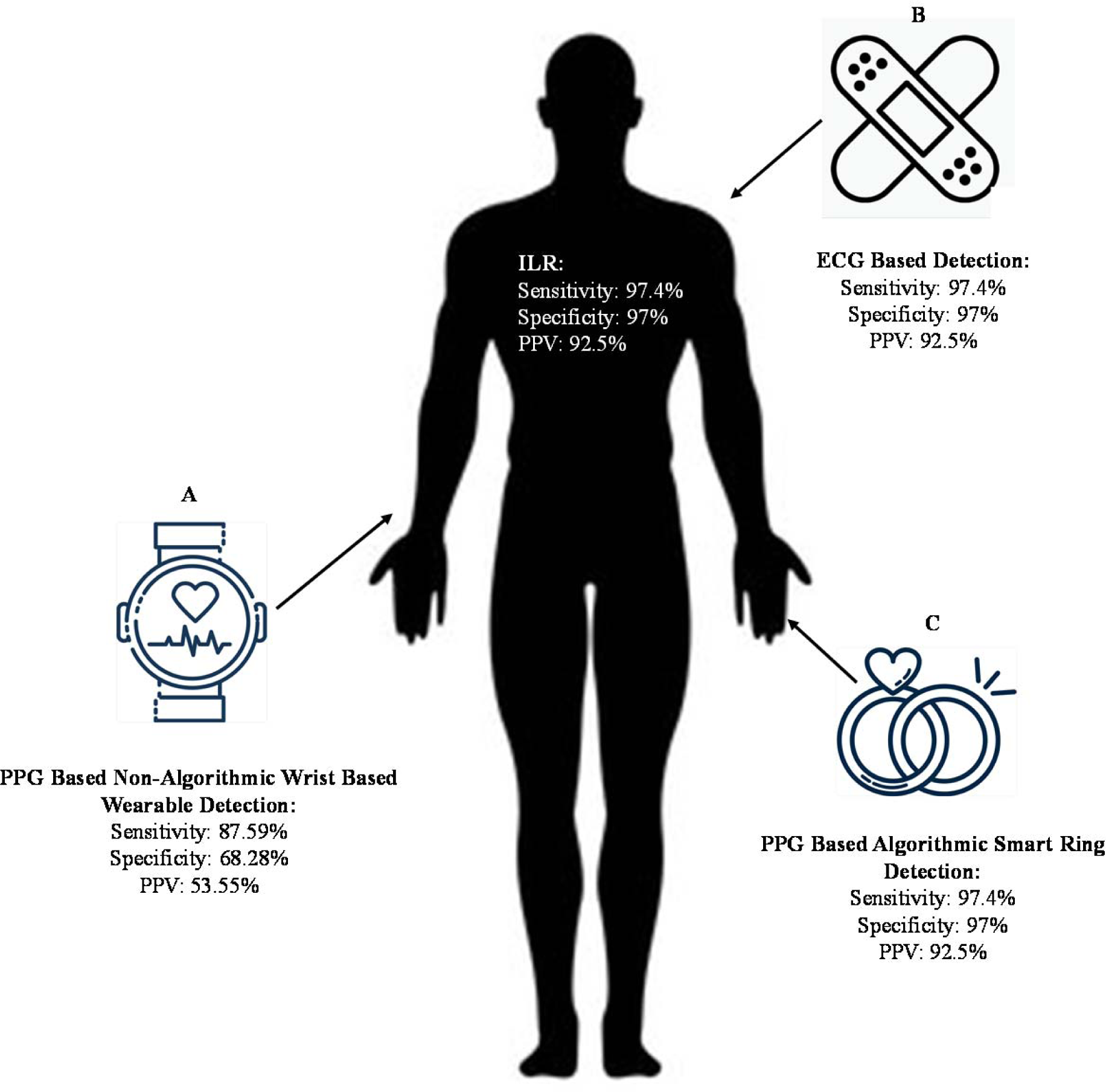
Illustration to show sensitivity, specificity and PPV of smart devices in detecting recurrence of atrial arrhythmias in post intervention patient cohorts compared with gold standard ILR compared to our data. **A.** REMOTE-AF data from 35 patients. **B.** ECG based detection post ablation in study of 99 patients.^24^. **C.** Smart ring-based detection of atrial arrhythmia post DCCV in study of 35 patients.^30^ ILR = implantable loop recorder. DCCV = direct current cardioversion.

Recurrence of paroxysmal and persistent atrial arrhythmias which are rate -controlled are likely to be misclassified as non-recurrence by our novel composite method. However, our NPV consistently being above 90% for all analysed groups shows that this had a minimal effect on our results. Our secondary endpoint showed that in patients with recurrence of atrial arrhythmia, patients described a poorer quality of life as evidenced by a higher AFEQT score when compared to the non-recurrence group. Consequently, our results demonstrate promise in identifying atrial arrythmia recurrence by combining PPG- recorded, non-exercise related elevations in heart rate with quality-of-life data. As a result, there is potential to avoid the invasive nature of ILR insertion as well as labour and cost intensive follow up to achieve long-term cost-effective monitoring. With further improvement in diagnostic accuracy, this could lead to an electronic point- of- care notification being sent via a smart device to assist pill in the pocket arrhythmia and anticoagulation management strategies as well as timely interventions (DCCV or redo catheter ablation) in patients experiencing recurrence of atrial arrythmias.^28, 29^ Lastly, due to our recruitment resulting in a homogenous population, more heterogeneity with regards to ethnicity and gender is required to draw more specific conclusions.

To our knowledge, our study is the first to utilise a novel PPG derived HR spike score methodology to detect AF/AT recurrence using gold- standard ILR technology as a comparator. The ILR (Reveal LINQ™) used in this study was found to have an AF detection sensitivity of 97.4%; specificity 97.0%; PPV 92.5%; NPV 99.0% and was able to detect AF burden with a sensitivity of 98.4%.^15^ Therefore, the ILR serves as an excellent comparator.

The use of mHealth wearables to detect atrial arrhythmia recurrence is gaining traction and to our knowledge REMOTE-AF is one of two studies, to report results for atrial arrythmia recurrence in patients with a known history of AF, utilising PPG enabled wearable technology. The other study utilised a smart ring to detect atrial arrythmia recurrence in a patient cohort post DCCV.^30^ Currently active is the SMART-ALERT clinical trial which will also use the ILR as a comparator for PPG -detected AF from a wearable device, both a smartwatch and smart ring. Data from this study can be used to further support or refute the use of PPG based wearables to detect atrial arrhythmias.^31^

Another study in progress is the SAFER (ISRCTN72104369)^32^ trial which will evaluate evidence to support the tolerability and feasibility of screening patients > 65 years old using PPG- enabled wearables. Our REMOTE-AF cohort with a mean age of 70.3 years and median follow up at 10 months, has already clearly demonstrated acceptability and tolerability in an older patient cohort. We recorded good compliance with device use and a roughly equal proportion of use during day-time and night-time hours. Alongside results reported by the eBRAVE-AF trial (NCT04250220)^29^ which screened for AF using smartphone PPG sensors, this leads to greater confidence that digital wearable technology can safely be used for not just screening but monitoring of known arrhythmia patients in the cohort at greatest risk of AF and its complications. Limited economic analyses demonstrate affordability and cost-savings associated with AF detection using wearables, specifically in over-65s, further highlighting their potential to improve health outcomes as measured by quality-adjusted-life-years by enhancing primary and primordial prevention.^33, 34^

Incorporating heart rate variability (HRV) into our novel composite method would likely have further improved our PPV as significant changes to HRV have been shown to correlate with AF.^35^ The Fitbit application programming interface (API) integrating HRV data was not open to researchers at the commencement of the study therefore REMOTE-AF study team did not have access to this data.

Future work requires more extensive investigation of PPG- enabled wearable devices and arrhythmia detection tools in real-world clinical settings with the design of larger trials including randomised clinical studies to report major adverse cardiovascular event (MACE) outcomes. A reduction in time to detection of AF/AT recurrence would allow for more responsive implementation of treatment strategies and lifestyle modifications to reduce disease burden and improve quality of life. Consumer mHealth devices can also act to further health literacy by facilitating patient understanding of disease progression and encourage positive behaviour change to modify their risk profile. As PPG technology continues to evolve, we can cautiously begin to consider this technology as an asset to improve clinical outcomes and guide treatment decisions.

We believe this proof- of- concept study can provide a foundation for further research and to develop this method in conjunction with sophisticated algorithmic development tools to inform a comprehensive diagnostic method to detect atrial arrhythmias with precision.

### Limitations

PPG technology itself has many limitations with the most prominent being motion artefact and noise interference, which can disrupt arrhythmia detection algorithms and signal quality, affecting the accuracy of heart rate measurements.^7, 36^ The devices used in the study were 2 years old with manufacturer recommended battery life of 4-5 days, however due to the lithium polymer battery life, gradual deterioration resulted in patients charging devices every 2-3 days which inevitably resulted in reduced compliance and more frequent interruptions to monitoring. The Fitbit Charge 2 is also only able to store data for 7 days before overwriting. To minimise data loss, we sent weekly text reminders to participants whose data did not synchronise in the past 7 days, however an automatic bi-weekly in-app notification would likely have increased adherence. A similar data upload issue was noted with the ILR. The Reveal LINQ™ device can store EGM data for 27 minutes of patient activated episodes and 30 minutes of automatic detected episodes,^37^ hence if multiple episodes are recorded over this time period without being downloaded to the base device, data is overwritten.

A further major limitation of the study was the small sample size and that all participants were Caucasian, potentially limiting the generalisability of our results. Sample size limitations prevented us from being able to power our study to derive statistical significance or draw wider conclusions applicable to a more diverse population. Given the exploratory nature of our study, our pilot data shows promise for further work to be undertaken with a sufficiently powered population size. Interruptions to ILR data upload via the CARELINK remote monitoring software is also acknowledged as a major limitation. Dependent on the frequency of arrhythmia recurrence, a more intensive download of ILR data would have been required to capture all EGM episodes of detected AF/AT recurrence (that are then able to be validated) rather than text only episodes. Our method of using text only episodes to confirm ILR detected AF/AT recurrence is likely to have affected the sensitivity of our results and is acknowledged as a significant limitation. In the two patients where we used pacemaker rhythm recording capabilities instead of ILR, we may have missed several episodes of AF/AT recurrence as episodes not documented on pacing reports at three monthly device interrogations had to be excluded. Furthermore, assessment of ILR EGMs was undertaken by a single reviewer therefore we recognise possible misclassifications may have occurred.

Finally, the research team did not have access to raw PPG waveform or HRV data which possibly led to a lower PPV. Applying the normalised spike score to raw PPG waveforms or incorporating HRV data into our novel composite method may have led to improvements in PPV.

Future studies that use a large variety of data modalities, similar to those recorded in REMOTE-AF, will need specific expertise in data handling and advanced data science due to the complexity of data acquired from wearables. With appropriate training and education for HCPs, it will enable a stepwise framework to incorporate high-value AI into routine cardiovascular care.^38^

## Conclusion

Our novel composite of wearable device data (PPG recorded heart rate data, HR spike score and step count) is a promising and modest predictor of AF/AT recurrence, in a post-ablation cohort compared to the gold standard ILR. Further work is required to determine whether consumer wearables integrating HR and step count with advanced algorithmic detection tools can improve AF/AT detection and guide treatment strategies.

## Data Availability

The data that support the findings of this study are available from the First Author [GA] upon reasonable request.

## Non Standard Abbreviations and Acronyms

AF: atrial fibrillation
AFEQT: atrial fibrillation effect on quality of life
AI: artificial intelligence
API: application programming interface
AT: atrial tachycardia
AWS: amazon web services
EHRA: european heart rhythm association
ESC: european society of cardiology
EGM: electrogram
HCP: healthcare professional
HR: heart rate
HRS: heart rhythm society
HRV: heart rate variability
ILR: implantable loop recorder
IQR: interquartile range
LSPAF: long standing persistent atrial fibrillation
mHealth: mobile health
NICE: national institute for health and care excellence
NPV: negative predictive value
PHE: public health england
PPG: photoplethysmography
PPV: positive predictive value
RADAR-base: remote assessment of disease and relapses
SD: standard deviation

## Acknowledgements

We would like to thank the CASA-AF research team at the Royal Brompton and Harefield Hospitals and the card*AI*c team at the University of Birmingham/University Hospitals Birmingham NHS Foundation Trust for set up of the study, as well as Grace Augustine (senior cardiac physiologist at Kettering General Hospital NHS Foundation Trust) for helping with the validation using ILR. We would also like to thank Julia Kurps, Joris Borgdorff and Nivethika Mahasivam at the Hyve (Utrecht, The Netherlands) for providing technology support, the RADAR-base team (Richard Dobson, Amos Folarin, Yatharth Ranjan, Pauline Conde, Callum Stewart and Zulqarnain Rashid at King’s College London, UK) and the BigData@Heart Consortium (Lead: Diederick E Grobbee, University Medical Centre Utrecht, The Netherlands; Scientific Coordinator: Folkert W Asselbergs, University of Amsterdam, The Netherlands).

